# A 5-min RNA preparation method for COVID-19 detection with RT-qPCR

**DOI:** 10.1101/2020.05.07.20055947

**Authors:** Alim Ladha, Julia Joung, Omar O. Abudayyeh, Jonathan S. Gootenberg, Feng Zhang

**Affiliations:** Department of Biological Engineering, Massachusetts Institute of Technology, Cambridge, MA 02139, USA; McGovern Institute for Brain Research at MIT; Howard Hughes Medical Institute, Cambridge, MA 02139, USA; Broad Institute of MIT and Harvard, Cambridge, MA 02142, USA; Department of Brain and Cognitive Sciences, Massachusetts Institute of Technology, Cambridge, MA 02139, USA

## Abstract

RNA extraction has become a bottleneck for detection of COVID-19, in part because of reagent shortages. We present here a rapid protocol that circumvents the need for RNA extraction that is compatible with RT-qPCR-based detection methods.

## A. Overview

The novel coronavirus SARS-CoV-2 causes COVID-19 and has resulted in an international public health emergency, spreading to over 180 countries and infecting more than 350,000 individuals. Testing for the presence of the virus is of utmost importance to both reduce the basic reproductive rate of the virus (R_0_) and inform best clinical practices for affected patients. However, understanding the full extent of the virus outbreak has remained challenging due to bottlenecks in the diagnosis of infection.

One of the major bottlenecks for COVID-19 diagnosis is the limited availability of RNA extraction kits for preparing virus RNA from patient samples and the low-throughput nature of the extraction procedure. Here, we describe an one-step column-free RNA preparation method that can be carried out in 5 minutes. The reaction can be used directly with the CDC COVID-19 RT-qPCR testing protocol, thus increasing throughput and alleviating supply chain issues.

## B. Materials and Reagents

- Quick Extract™ DNA Extraction Solution ([QE09050](https://www.lucigen.com/QuickExtract-DNA-Extraction-Solution/)), Lucigen. Once thawed, aliquot and store at −20C to avoid >3 freeze-thaw cycles.

## C. Protocol

1. Dilute nasopharyngeal swab stored in Viral Transport Medium or Human Specimen Control (HSC) 1:1 with Quick Extract™ DNA Extraction Solution. For example, in a fresh PCR tube, mix 20 µl of swab sample with 20 µl of Quick Extract.
2. Incubate swab and Quick Extract mix at **95°C for 5 minutes**. Allow reaction to cool on ice before proceeding.
3. Use reaction from step (2) for qRT-PCR. Use an amount from step (2) that corresponds to 10 % of the total qRT-PCR reaction volume. For example, for a reaction with total volume of 50 µl, use 5 µl of the reaction from step (2).

## D. Assay Development and Preliminary Validation

We evaluated a number of buffer compositions to identify one that achieved efficient lysis of enveloped virus while preserving the activity of the CDC recommended RT-qPCR reaction (TaqPath™ 1-Step RT-qPCR Master Mix). Of all of the buffers tested, Quick Extract™ DNA Extraction Solution provided results that were comparable to standard RNA extraction kits.

To confirm that the presence of QE does not interfere with RT-qPCR activity, we compared RT-qPCR reactions using synthetic SARS-CoV-2 gene fragment (Twist Synthetic SARS-CoV-2 RNA Control 1, SKU:102019) dissolved in either ddH_2_O or in a 50:50 ddH_2_O:Quick Extract mixture. We set up each RT-qPCR reaction with a total volume of 10 µl (1 µl of RNA sample, 0.5 µl of CDC probe N1, 2.5 µl of TaqPath RT-qPCR master mix, and 6 µl of ddH_2_O). From these reactions, we found that Quick Extract at a final concentration of 5 % did not negatively affect the RT-qPCR reaction (Figure 1A; 2 technical replicates).

**Figure 1.**
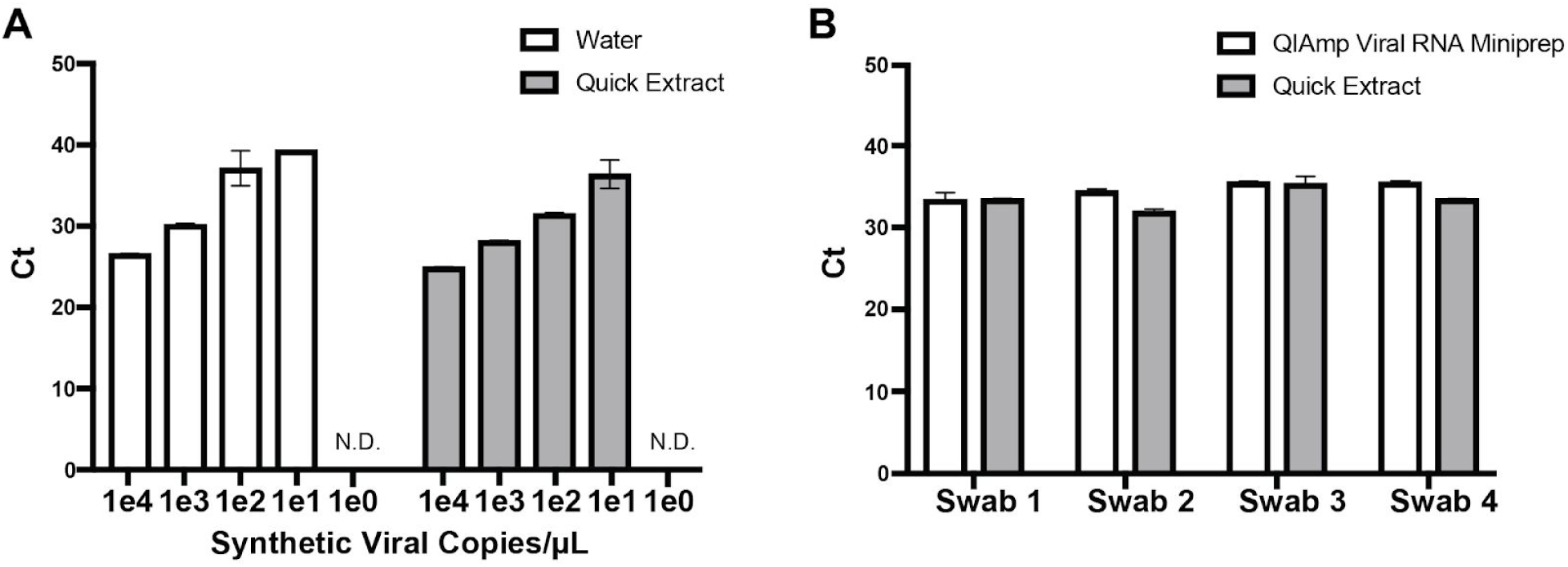
**A)** RT-qPCR of synthetic SARS-CoV-2 RNA control (Twist Synthetic SARS-CoV-2 RNA Control 1, SKU:102019) diluted in water or 50:50 ddH_2_O:Quick Extract mixture. Final sample volume was 1 µL in a 10 µL reaction. **B)** RT-qPCR of diluted COVID-19 positive nasopharyngeal swabs treated at 95°C for 5 minutes in Quick Extract or purified using QIAmp Viral RNA Miniprep.

We conducted preliminary validation of the Quick Extract RNA preparation procedure on COVID-19 positive nasopharyngeal swabs and found that RNA samples prepared using Quick Extract supported similarly sensitive detection of coronavirus as QIAmp Viral RNA Miniprep (Qiagen) for all 4 swab samples (Figure 1B; 2 biological replicates). To simulate low viral load, coronavirus positive swabs were diluted 1:10 in pooled nasopharyngeal swabs from 5 healthy donors ([Lee Biosolutions, SKU:991-31-NC-5](https://www.leebio.com/product/245/nasal-cavity-swab-human-donor-991-31-nc)) prior to purification or Quick Extract treatment. For the QIAmp Viral RNA Miniprep conditions, 100 µL of diluted swab sample was used for extraction and was eluted using 100 µL of ddH_2_O. 1 µL of the elution was used in a 10 µL RT-qPCR reaction. For the Quick Extract conditions, 1 µL of Quick Extract preparation was used for each 10 µL RT-qPCR reaction.

## Data Availability

All of the data is presented in the paper.

## Acknowledgements

We would like to thank Keith Jerome, Alex Greninger, Meei-Li Huang, and Bob Bruneau of the University of Washington for generously providing COVID-19 nasopharyngeal swab samples. We would like to acknowledge support from the McGovern Institute; the Howard Hughes Medical Institute; Open Philanthropy Project; James and Patricia Poitras; and Robert Metcalfe.

## Ethical Statement

Patient samples were existing biospecimens and de-identified. This work (E-2031, CRISPR-based Diagnostics) was determined to be “not human subjects research” by MIT (as defined by Federal regulation 45 CFR 46 under the following: Exempt Category 4 - Secondary Use Research).

